# Vaginal-Stimulating Product Use and Cervicovaginal Health Among Adolescent Girls and Young Women in Rural South Africa: A Cross-Sectional Baseline Study

**DOI:** 10.64898/2026.03.23.26349137

**Authors:** Phumla Radebe, Sibeko Singeziwe, Maphumulo Nokuthula, Masson Lindi, Humphries Hilton, Mntambo Ntombenhle, Ndlela Nonsikelelo, Liebenberg Lenine, Ngcapu Sinaye, Samsunder Natasha, Potloane Disepo, Saruchera Bester, Abdool Karim Quarraisha, Jaspan Heather, Passmore Jo-Ann, Mkhize Pamela

## Abstract

**Background:** Vagina-stimulating products (VSPs) are widely used in sub-Saharan Africa for perceived sexual enhancement, hygiene, or cultural practices, yet their biological implications for cervical health remain poorly understood, particularly among adolescent girls and young women (AGYW). These practices occur within largely unregulated informal markets and intersect with gender norms, sexual negotiation, and reproductive health vulnerability among AGYW. We present baseline findings from a prospective cohort examining associations between VSP use, cervicovaginal abnormalities, and sexually transmitted infections (STIs).

**Methods:** Cross-sectional baseline data were analysed from 252 sexually active, HIV-negative participants enrolled in rural KwaZulu-Natal, including adolescents (n=188, 14–19 years) and adults (n=64, 25–35 years). Participants completed structured questionnaires and underwent clinical examination, STI testing, human papillomavirus (HPV) genotyping, vaginal pH assessment, and colposcopy. VSP use was categorised as intravaginal, ingested, or none. Age-stratified multivariable logistic regression models estimated statistical associations, adjusting a priori for age, hormonal contraceptive use, condom use, vaginal pH and lifetime number of sexual partners.

**Results:** VSP use was common among adolescents (68.1%) and adults (69.8%), with adolescents more likely to report intravaginal use and adults more likely to report ingested products. Among adolescents, VSP use was not statistically associated with cervicovaginal abnormalities after adjustment. However, exploratory analyses showed higher prevalence of *Trichomonas vaginalis* infection (p=0.021) and HPV-16 (p=0.007) among VSP users. Alum use was statistically associated with lower odds of visible cervicovaginal injury (OR 0.14; 95% CI 0.04–0.48), though this finding should be interpreted cautiously. Among adults, both intravaginal (aOR 3.7; 95% CI 1.2–11.8) and ingested (aOR 2.9; 95% CI 1.1–8.5) VSP use were statistically associated with cervical ectopy, while product-specific associations with individual products did not persist after adjustment.

**Conclusion:** VSP use is highly prevalent among AGYW, with distinct age-specific patterns. In this cross-sectional baseline analysis, VSP use was statistically associated with cervical ectopy among adults, while exploratory findings suggested associations with STI and HPV patterns among adolescents. These hypothesis-generating findings underscore the need for longitudinal and mechanistic studies to clarify biological pathways linking VSP use with infection risk, and highlight the potential importance of culturally responsive sexual health interventions and regulatory attention to informal vaginal product markets.

## Introduction

Sexually transmitted infections (STIs), including human immunodeficiency virus (HIV) and human papillomavirus (HPV), remain major global public health challenges and disproportionately affect women in sub-Saharan Africa (SSA). Persistent infection with high-risk (HR) HPV types is the necessary cause of cervical cancer (1), while curable STIs such as *Chlamydia trachomatis* (CT), *Neisseria gonorrhoeae* (NG), and *Trichomonas vaginalis* (TV) continue to contribute substantially to reproductive morbidity worldwide (3,4). Despite global commitments to cervical cancer elimination and STI control, behavioural and structural practices that may alter mucosal vulnerability remain under-characterised in high-burden settings.

Adolescent girls and young women (AGYW) in SSA, particularly those aged 14–35 years in South Africa (SA), bear a disproportionate burden of HIV, HPV, and other STIs (4). Despite the scale-up of preventive interventions, including condom promotion, pre- and post-exposure prophylaxis (PrEP and PEP), and expanded HIV testing, STI incidence remains high in this population (6). This persistent burden has prompted increased interest in identifying biological and behavioural factors that may influence infection acquisition and persistence among AGYW (4,7–9).

One set of practices that may influence cervicovaginal health, yet remains insufficiently characterised biologically, is the use of vagina-stimulating products (VSPs). These products are widely used across SSA for perceived sexual enhancement, hygiene, or cultural purposes. They include intravaginal and ingested formulations intended to alter the vaginal environment (10–14). Social and commercial narratives promoting vaginal tightness, dryness, and cleanliness may further reinforce these practices (15,16). Unlike regulated pharmaceutical or cosmetic products, many VSPs are informally produced and distributed, and their chemical composition is often unknown.

Qualitative research from rural, high-HIV-burden KwaZulu-Natal has highlighted the socio-relational motivations underpinning VSP use, particularly among adolescents, including partner satisfaction, conformity to social norms, and negotiation of gendered power dynamics (12). While these practices are socially meaningful, experimental and observational studies suggest that some intravaginal products may alter mucosal integrity, induce epithelial irritation, or modify local immune responses (13,17,18). Such effects have been hypothesized to influence susceptibility to infection, although direct clinical evidence remains limited. Existing literature has largely focused on behavioural associations or self-reported symptoms, with relatively few studies integrating colposcopic examination and molecular diagnostics.

To address these gaps, we conducted a cross-sectional baseline analysis within a prospective cohort of AGYW in KwaZulu-Natal, South Africa. Using structured questionnaires alongside colposcopy, STI testing, and HPV genotyping, we aimed to: 1) describe the prevalence and patterns of VSP use; 2) compare cervicovaginal abnormalities between VSP users and non-users; and 3) examine associations between VSP use and selected STIs (TV, CT, NG, and HPV), as well as other vaginal infections including bacterial vaginosis (BV) and vulvovaginal candidiasis. These baseline findings provide clinically contextualised and molecularly characterised data to inform longitudinal analyses of VSP exposure, mucosal integrity, infection susceptibility, and potential implications for STI and cervical cancer prevention strategies.

## Methods and Materials

### Study design

This analysis used baseline data from the Mucosal Injury from Sexual Contact (MISC) prospective cohort study conducted at the Centre for the AIDS Programme of Research in South Africa (CAPRISA). The MISC study investigates socio-behavioural, anatomical, and biological factors influencing cervicovaginal health following vaginal sex and/or vaginal-stimulating product (VSP) use among adolescent (14–19 years) and adult (25–35 years) women. For the present analysis, only enrolment (baseline) data were included. Data on VSP use, sexual behaviour, and clinical outcomes, including cervicovaginal abnormalities, sexually transmitted infections (STIs), and other vaginal infections, were collected using structured questionnaires, colposcopy, and molecular assays. As analyses were limited to baseline observations, findings are cross-sectional and do not imply causality. This study was reported in accordance with the STROBE guidelines for observational studies

### Study setting

The study was conducted at the rural CAPRISA Vulindlela research clinical site in the uMgungundlovu district, outside of Durban, KZN, SA. The area is characterised by socio-economic challenges, including limited infrastructure, as well as a high prevalence of STIs and HIV (19).

### Study population and sample size

A total of 353 women without HIV were prospectively recruited between 21/11/2017 and 10/11/2021 as part of the study cohort. Participants completed demographic, reproductive health, behavioural, and VSP-use questionnaires. Of these, 252 participants had complete baseline clinical and laboratory data and were included in the cross-sectional analysis. The remaining 101 participants were excluded because of incomplete baseline data, ineligibility, or withdrawal prior to completion of enrolment procedures (S1 Fig, S1 Table). Participants were enrolled irrespective of VSP exposure status; exposure was determined based on self-reported use at baseline.

### Ethical considerations

Ethical approval was obtained from the Biomedical Research Ethics Committee of the University of KwaZulu-Natal (BREC/00003306/2021) and the Institutional Review Board of Seattle Children’s Hospital (STUDY00000462). All participants provided written informed consent prior to enrolment. Adolescents aged 14–17 years provided written parental or guardian consent and participant assent which were obtained in accordance with ethical guidelines.

### Clinical procedures

Eligible participants were sexually active, living without HIV, non-pregnant, had no history of cervical disease, had not used antibiotics in the preceding four weeks, and were able to provide informed consent (or assent with parental consent for minors). Participants were instructed to abstain from VSP use and sexual intercourse for at least two weeks prior to the baseline visit to allow mucosal recovery. Adherence was assessed via structured questionnaire at the study visit; no biological verification of abstinence was performed.

### Colposcopy examination and assessment of mucosal trauma

Cervical examinations were conducted by qualified nurses trained in study-specific procedures using a hand-held EVA System colposcope (MobileODT, Tel Aviv, Israel). Digital images were independently reviewed by the study gynaecologist. Completed colposcopy records were available for 180 adolescents and 58 adults and were included in this analysis.

Cervical ectopy was defined as the extension of endocervical columnar epithelium onto the ectocervix and graded according to the proportion of the ectocervical surface involved (≤25%, ≤50%, ≤75%, or >75%), following validated criteria (21). Other cervicovaginal abnormalities were assessed using the Manual for the Standardization of Colposcopy for the Evaluation of Vaginal Products (22) and included leukoplakia, mucosal trauma (petechiae, ecchymosis, lacerations, abrasions, or bleeding), inflammation (erythema, oedema, or mucosal peeling), genital warts, and abnormal vaginal discharge.

Cervicovaginal trauma included findings such as petechiae, bleeding, ecchymosis, lacerations, cervical abrasions, or the presence of blood clots on the cervix. Inflammation, which may overlap with injury-related findings, was defined as the presence of erythema, oedema, or mucosal peeling. Grossly white findings (leukoplakia) were identified as flat, opaque white plaques occasionally exhibiting slight ridging or surface corrugation. Genital warts (condyloma acuminata) and abnormal vaginal discharge (frothy, white, creamy, brownish, clear, or greenish, with or without odour) were also recorded.

### Specimen collection and STI testing

Urine specimens were collected for pregnancy testing prior to genital specimen collection. Urine was tested for *Chlamydia trachomatis* (CT) and *Neisseria gonorrhoeae* (NG) using the Cepheid Xpert CT/NG assay (GeneXpert® Instrument System). Vaginal swabs obtained from the lateral vaginal wall or posterior fornix were used to assess bacterial vaginosis (BV) by Nugent scoring and *Trichomonas vaginalis* (TV) infection using the OSOM® Trichomonas Rapid Test and wet mount microscopy. Potassium hydroxide microscopy was performed to detect fungal hyphae or yeast. Vaginal pH was measured using pH test strips and recorded using a reference chart.

### Human papillomavirus (HPV) genotyping

Endocervical swab specimens eluted in phosphate-buffered saline were centrifuged, and the pellet resuspended in DNase/RNase-free water for cell lysis. Aliquots of lysate were amplified using the HPV Direct Flow Chip Kit (Master Diagnóstica, Spain) according to the manufacturer’s protocol. Amplified products were hybridised onto membrane chips containing genotype-specific probes and analysed using the hybriSpot platform with hybriSoft software (version 2.0). The assay detects multiple low-risk (LR) and high-risk (HR) HPV genotypes.

### Statistical analysis

Analyses were stratified by age group (adolescent vs. adult) to account for age-related differences in sexual behaviour, cervicovaginal abnormalities, and STI prevalence. Categorical variables were compared using the chi-squared or Fisher’s exact test, as appropriate, and continuous variables were compared using the Wilcoxon rank-sum test. Univariable analyses explored associations between VSP use, genital infections, and cervical abnormalities. Multivariable logistic regression models were constructed to estimate adjusted associations while controlling for potential confounders selected a priori based on biological plausibility and prior literature, including age, hormonal contraceptive use, condom use, vaginal pH, and lifetime number of sexual partners. A two-sided p-value <0.05 was considered statistically significant, and 95% confidence intervals (CIs) are reported. Analyses were performed using GraphPad Prism (versions 8.4.3 and 9.4.1) and MATLAB R2024b.

## Results

### Participant characteristics

A total of 252 participants were enrolled, including 188 adolescents and 64 adults (S1Fig).

VSP use was commonly reported in both age groups (68.1% of adolescents; 69.8% of adults). Among adolescents, demographic and reproductive characteristics were similar between VSP users and non-users (Table 1).

**Table 1.** Demographic and reproductive health characteristics of adolescent VSP nonusers versus users.

| Characteristics | Non-user<br>[n (%)] | VP<br>[n (%)] | ± OR (95% CI) | Ingested<br>[n (%)] | ± OR (95% CI) |
| --- | --- | --- | --- | --- | --- |
| N | 63 | 78 |  | 47 |  |
| **<sup>a</sup>Age** |  |  |  |  |  |
| 14-19 Years | 18 [17 – 18] | 17 [16 – 18] | 0.6 (0.28 – 1.14) | 18 [17 – 18] | 0.56 (0.28 – 1.14) |
| **Highest educational level** |  |  |  |  |  |
| Primary school | 2/63 (3.2%) | 0/78 (0.0%) | 0.0 (0.0 – 1.74) | 2/47 (4.3%) | 1.4 (0.21 – 8.88) |
| Secondary school | 61/63 (96.8%) | 76/78 (97.4%) | 0.8 (0.12 – 5.25) | 2/47 (4.3%) | **686.3 (91.2 – 3505.0)** |
| Tertiary school | 0/63 (0.0%) | 2/78 (2.6%) | 0.0 (0.0 – 2.67) | 43/47 (91.5%) | **0.0 (0.0 – 0.01)** |
| **Employment status** |  |  |  |  |  |
| Employed | - | - | - | - | - |
| Unemployed | 63/63 (100.0%) | 78/78 (100.0%) | - | 47/47 (100.0%) | - |
| **Relationship status** |  |  |  |  |  |
| Living with partner | 1/63 (1.6%) | 2/78 (2.6%) | 0.6 (0.04 – 5.38) | 0/47 (0.0%) | 0.0 (0.00 – 12.06) |
| Living separately with a partner± | 62/63 (98.4%) | 76/78 (97.4%) | 0.6 (0.04 – 5.38) | 47/47 (100.0%) | 0.0 (0.0 – 12.06) |
| **Ever pregnant** |  |  |  |  |  |
| Yes | 19/63 (30.2%) | 18/78 (23.1%) | 0.6 (0.33 – 1.46) | 15/47 (31.9%) | 1.1 (0.48 – 2.36) |
| No | 44/63 (69.8%) | 60/78 (76.9%) |  | 32/47 (68.1%) |  |
| **Hormone Contraception use~** |  |  |  |  |  |
| Yes | 19/63 (30.2%) | 31/75 (41.3%) | 1.6 (0.81 – 3.21) | 13/47 (27.7%) | 0.9 (0.37 – 1.97) |
| Nuristerate | 9/63 (14.3%) | 12/75 (16.0%) | 0.9 (0.37 – 2.18) | 6/47 (12.8%) | 0.9 (0.28 – 2.65) |
| Depot | 5/63 (7.9%) | 3/75 (4.0%) | 0.5 (0.12 – 1.9) | 6/47 (12.8%) | 1.7 (0.45 – 5.34) |
| <sup>a</sup>Vaginal pH | 4.9 [4.7 – 5.3] | 4.7 [4.4 – 5.0] |  | 4.7 [4.4 – 5.0] |  |

<sup>a</sup> Numerical category reported as median [IQR]
± Odds ratio compares with adolescents or adult reporting to not using any products, calculated using the Chi-squared test
VP- Vaginally inserted product, Ingested- Oral products, Non-user- No product used

Hormonal contraceptive use was more frequently reported among VSP users than non-users (41.3% vs. 30.2%), although this difference was not statistically significant (OR 1.6; 95% CI 0.81–3.21). Median vaginal pH was slightly lower among VSP users than non-users (4.7 vs. 4.9). Educational level, employment status, relationship status, and pregnancy history did not differ by product use. Among adults, ingested products were the most commonly reported VSPs (44.4%), followed by intravaginal products (25.4%). Relationship status differed by product use; living with a partner was more frequently reported among intravaginal product users than non-users (25% vs. 0%; Table 2).

**Table 2.** Demographic and reproductive health characteristics of adult VSP nonusers and users.

| Characteristics | Non-user<br>[n (%)] | VP<br>[n (%)] | ± OR (95% CI) | Ingested<br>[n (%)] | ± OR (95% CI) |
| --- | --- | --- | --- | --- | --- |
| N | 19 | 16 |  | 26 |  |
| **<sup>a</sup>Age** |  |  |  |  |  |
| 25-35 Years | 28 [26.5 – 30.0] | 30 [27.0 – 32.0] | 1.5 (0.74 – 3.19) | 28 [26.0 – 30.0] | 1.54 (0.74 – 3.19) |
| **Highest educational level** |  |  |  |  |  |
| Primary school | 0/19 (0.0%) | 1/16 (6.3%) | 0.0 (0.00 – 7.58) | 26/28 (92.9%) | 0.0 (0.00 – 0.03) |
| Secondary school | 18/19 (94.7%) | 15/16 (93.8%) | 0.8 (0.04 – 16.79) | 2/28 (7.1%) | **0.0 (0.00 – 0.04)** |
| Tertiary school | 1/19 (5.3%) | 0/16 (0.0%) | 0.0 (0.00 – 10.69) | 0/28 (0.0%) | 0.0 (0.00 – 1.51) |
| **Employment status** |  |  |  |  |  |
| Employed | 4/19 (21.1%) | 1/16 (6.3%) | 0.3 (0.02 – 1.94) | 3/28 (10.7%) | 0.45 (0.10 – 1.90) |
| Unemployed | 15/19 (78.95%) | 15/16 (93.8%) | 4.0 (0.52 – 51.73) | 25/28 (89.3%) | 2.2 (0.53 – 9.61) |
| **Relationship status** |  |  |  |  |  |
| Living with partner | 0/19 (0.0%) | 4/16 (25.0%) | **0.0 (0.00 – 0.83)** | 1/28 (3.6%) | 0.0 (0.00 – 13.26) |
| Living separately with a partner | 19/19 (100.0%) | 12/16 (75.0%) | **0.0 (0.00 – 0.83)** | 27/28 (96.4%) | 0.0 (0.00 – 13.26) |
| **Ever pregnant** |  |  |  |  |  |
| Yes | 18/19 (94.7%) | 14/16 (87.5%) | 0.4 (0.02 – 3.70) | 24/28 (85.7%) | 0.3 (0.03 – 2.41) |
| No | 1/19 (5.3%) | 2/16 (12.5%) | 2.6 (0.27 – 39.04) | 4/28 (14.3%) | 3.0 (0.42 – 38.49) |
| **Hormone Contraception use~** |  |  |  |  |  |
| Yes | 8/19 (42.1%) | 6/16 (37.5%) | 0.8 (0.24 – 3.53) | 7/28 (25.0%) | 0.4 (0.12 – 1.55) |
| Nuristerate | 1/19 (5.3%) | 1/16 (6.3%) | 1.2 (0.06 – 23.93) | 3/28 (10.7%) | 2.2 (0.30 – 29.43) |
| Depot | 6/19 (31.6%) | 5/16 (31.3%) | 0.9 (0.23 – 3.79) | 9/28 (32.1%) | 1.0 (0.31 – 3.4) |
| <sup>a</sup>Vaginal pH | 4.7 [4.7 – 5.3] | 5.0 [4.7 – 5.3] |  | 4.7 [5.0 – 5.0] |  |

<sup>a</sup> Numerical category reported as median [IQR]
<sup>\*</sup> Odd ratio compares with adolescents or adult reporting to not using any products, calculated using the Chi-squared test VP- Vaginally inserted product, Ingested- Oral products, Non-user- No product used

### Patterns of vaginal-stimulating product use

Participants reported 46 unique products, including 27 intravaginal and 19 ingested (S2 Fig). Products included powders, crystals, gels, liquids, paper, and mixed traditional herbal preparations, and were used for vaginal heating, tightening, sexual stimulation, or hygiene/detoxification purposes. Adolescents were more likely than adults to report intravaginal product use (41.5% vs. 25.4%; p = 0.024), whereas adults were more likely to report ingested product use (44.4% vs. 25.0%; p = 0.007). Intravaginal alum use was more frequently reported among adolescents than adults (33.3% vs. 6.3%; p = 0.056). The use of ibhodwe labafazi (direct translation: “the women’s pot”), a pink-scented petroleum jelly of unknown contents, was more commonly reported among adults than adolescents (56.3% vs. 5.0%; p = 0.027).

### Cervical abnormalities observed on colposcopy by age group

Colposcopic evaluation was completed for 176 adolescents and 50 adults (Fig 1; S2 Table). Adults had a higher prevalence of visible cervicovaginal injury or inflammation than adolescents (28.0% vs. 5.7%; p < 0.001). Leukoplakia was the most frequently observed finding among adolescents and was less common among adults (48.9% vs. 30.0%; p = 0.018). Cervical ectopy was observed in 4.5% of adolescents and 12.0% of adults, with a non-significant trend toward higher prevalence among adults (p = 0.054). The prevalence of genital warts and abnormal vaginal discharge did not differ significantly by age group.

**Fig 1.**
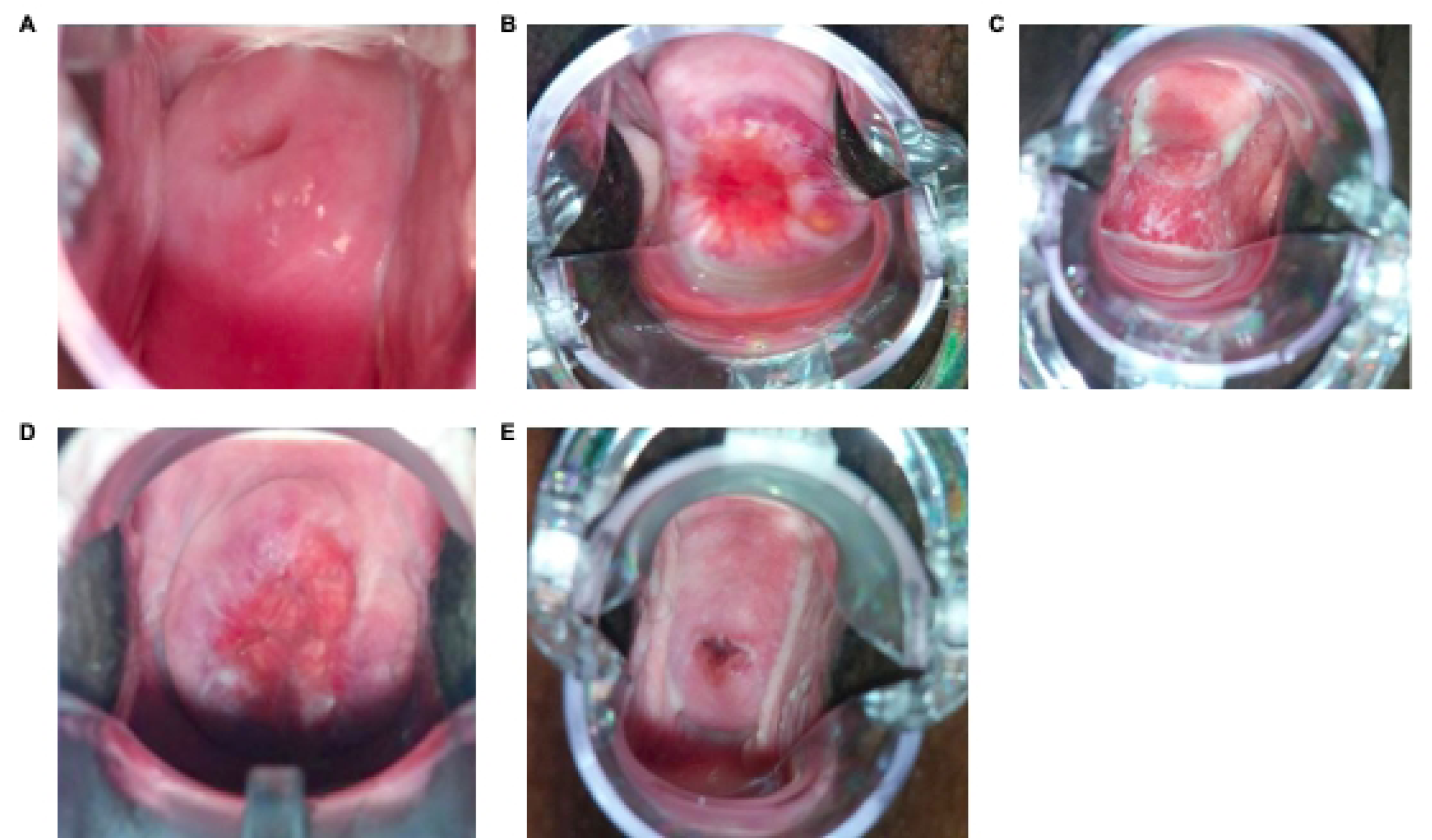
Description of cervical abnormalities included in the study. Colposcopy images were captured using the EVA system colposcope. The studied cervical abnormalities were classified as, A) Normal cervix; B) Cervical ectopy; C) and D) which were signs of injury indicated by: C) petechiae and D) erythema, or clinically evident inflammation; E) Grossly white findings – Leukoplakia. Genital warts and vaginal discharge were also analysed (images not shown).

### Associations between VSP use and cervicovaginal abnormalities

Associations between VSP use and cervicovaginal abnormalities were examined separately by age group (Table 3). Among adolescents, neither intravaginal nor ingested VSP use was significantly associated with cervical ectopy, leukoplakia, visible injury/inflammation, genital warts, or abnormal vaginal discharge in unadjusted analyses. Among adults, both intravaginal and ingested VSP use were significantly associated with cervical ectopy. Cervical ectopy was observed in 35.3% of intravaginal product users compared to 5.3% of non-users (p = 0.023) and in 31.8% of ingested product users compared to non-users (p = 0.032). No significant associations were observed between adult VSP use and leukoplakia, injury/inflammation, or abnormal vaginal discharge.

**Table 3.** Comparison of cervico vaginal colposcopic findings amongst vaginal product users and non-users in KZN at baseline.

| Variable | No product(s)<br>[% (n/N)] | Vaginal product<br>user<br>[% (n/N)] | *Unadjusted P-value | Oral product<br>user<br>[% (n/N)] | *Unadjusted<br>value |
| --- | --- | --- | --- | --- | --- |
| **Baseline visit Adolescents (N=156)** |  |  |  |  |  |
| Cervical ectopy | 10.3% (6/58) | 15.4% (10/65) | 0.407 | 12.1% (4/33) | 0.795 |
| Leukoplakia | 51.7% (30/58) | 40.0% (26/65) | 0.479 | 42.4% (14/33) | 0.211 |
| Injury | 8.6% (5/58) | 7.7% (5/65) | 0.765 | 18.2% (6/33) | 0.179 |
| Genital warts | 3.4% (2/58) | 1.5% (1/65) | 0.456 | 0.0% (0/33) | 0.281 |
| Vaginal discharge | 25.9% (15/58) | 35.4% (23/65) | 0.234 | 27.3% (9/33) | 0.883 |
| **Baseline visit Adults (N=58)** |  |  |  |  |  |
| Cervical ectopy | 5.3% (1/19) | 35.3% (6/17) | **0.023** | 31.8% (7/22) | **0.032** |
| Leukoplakia | 47.4% (9/19) | 23.5% (4/17) | 0.137 | 27.3% (6/22) | 0.183 |
| Injury | 26.3% (5/19) | 23.5% (4/17) | 0.847 | 36.4% (8/22) | 0.491 |
| Genital warts | - | - |  | - | - |
| Vaginal discharge | 21.1% (4/19) | 17.6% (3/17) | 0.865 | 4.5% (1/22) | 0.107 |

p-value <0.05 is considered significant and unadjusted p-values were calculated using the \*Chi-squared test
Cervicovaginal colposcopic findings were grouped into: Leukoplakia; Injury- erythema, peeling, edema, ecchymosis, laceration, and abrasion.

### Product-specific associations with cervicovaginal abnormalities

Product-specific regression analyses were conducted for commonly reported products (alum, ibhodwe labafazi, snuff, and other vaginal products; Fig 2; Fig 3).

**Fig 2.**
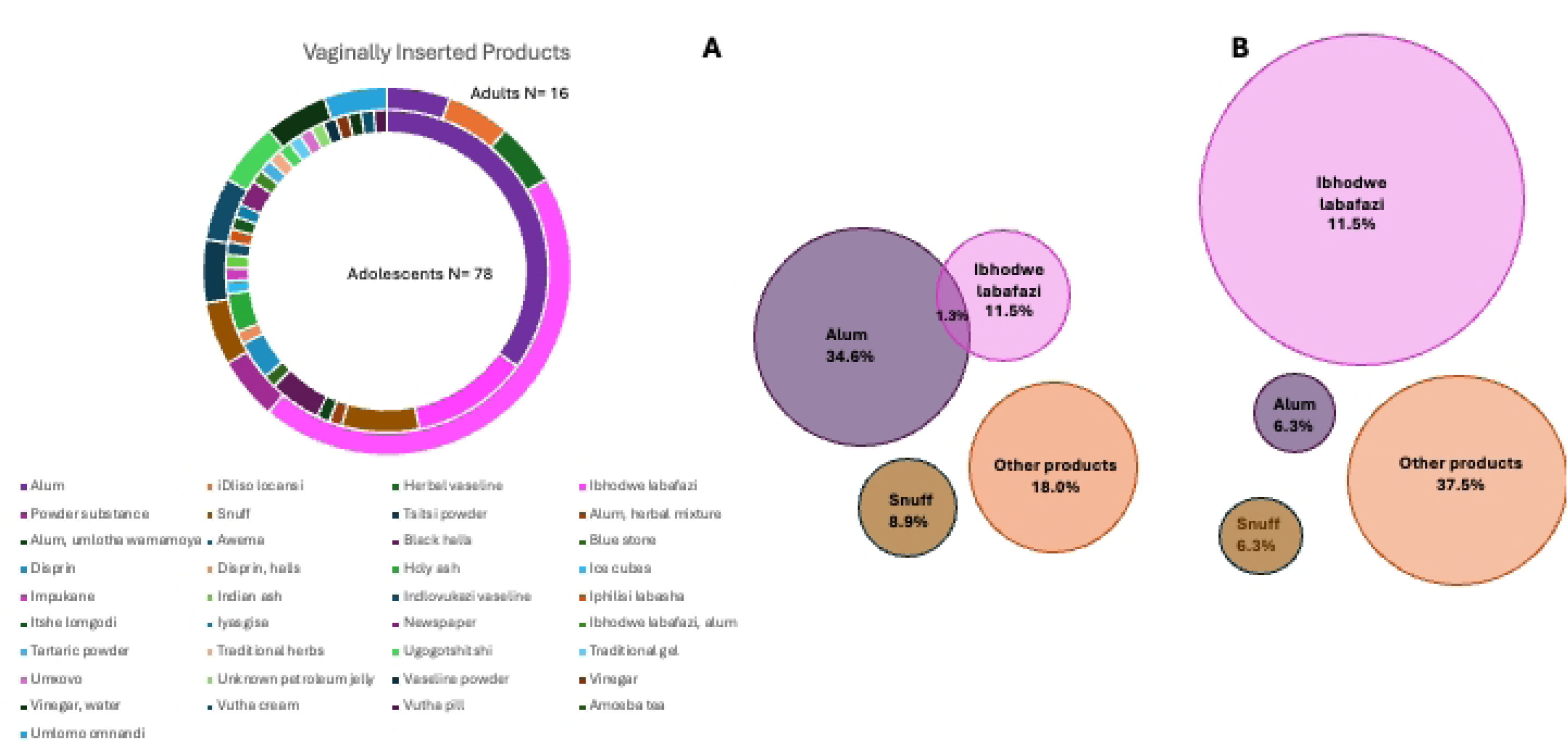
Shared product use by adolescent female vs adult women. The area proportional Venn diagram presents the proportions of the most exclusive intravaginal products used by women in KZN- A) Adolescents alum (purple), ibhodwe labafazi (pink), snuff (brown); B) Adults ibhodwe labafazi (pink), alum (purple), snuff (brown). Among adolescents, alum use was statistically associated with lower odds of visible cervicovaginal injury or inflammation compared with non-use (OR 0.14; 95% CI 0.04–0.48; p = 0.002; Fig 3A). A non-significant trend toward increased injury or inflammation was observed among adolescents reporting intravaginal snuff use. No associations were observed between adolescent VSP use and leukoplakia, cervical ectopy, or abnormal vaginal discharge after adjustment for age,condom use, hormonal contraceptive use, and lifetime sexual partners.

**Fig 3.**
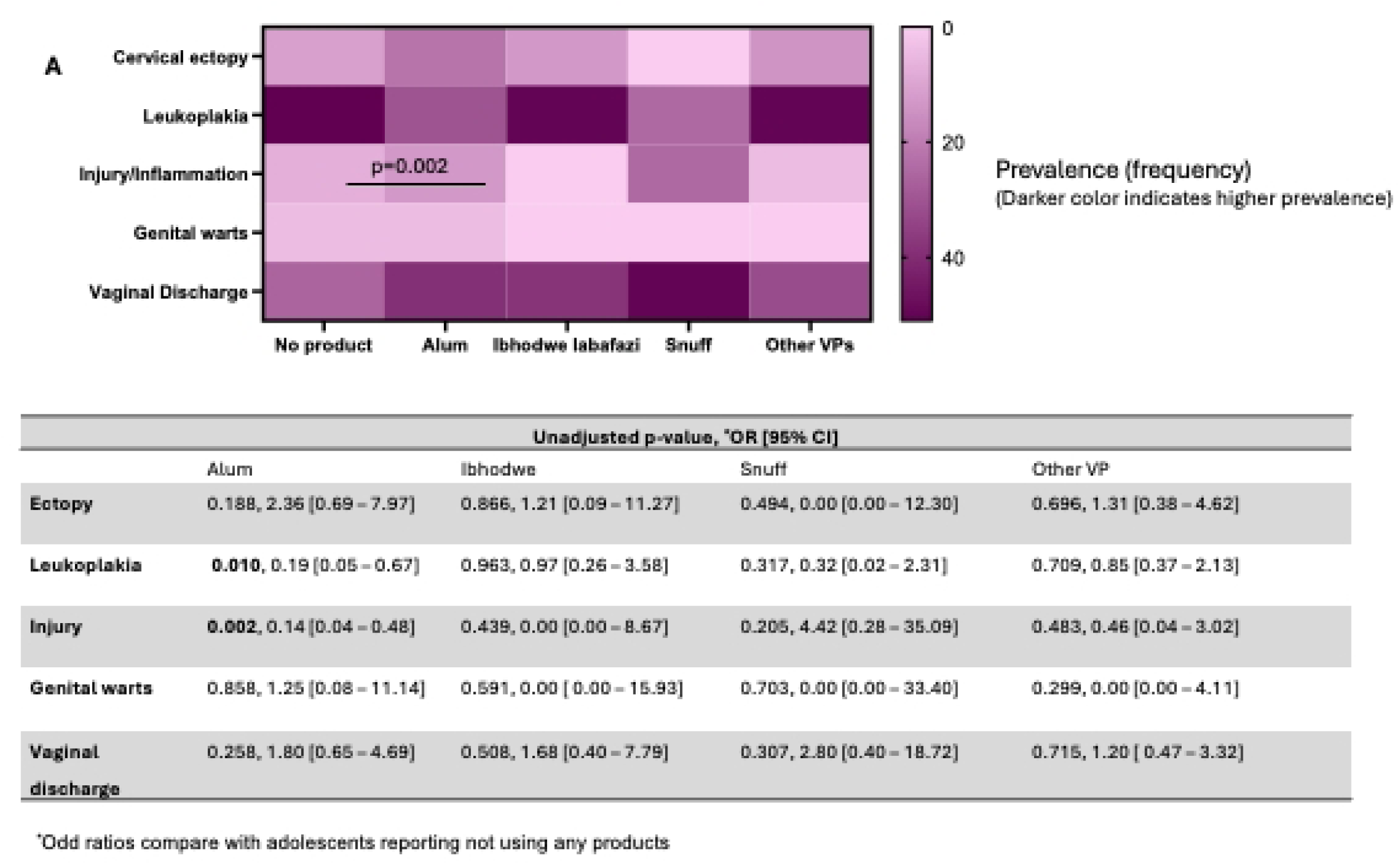
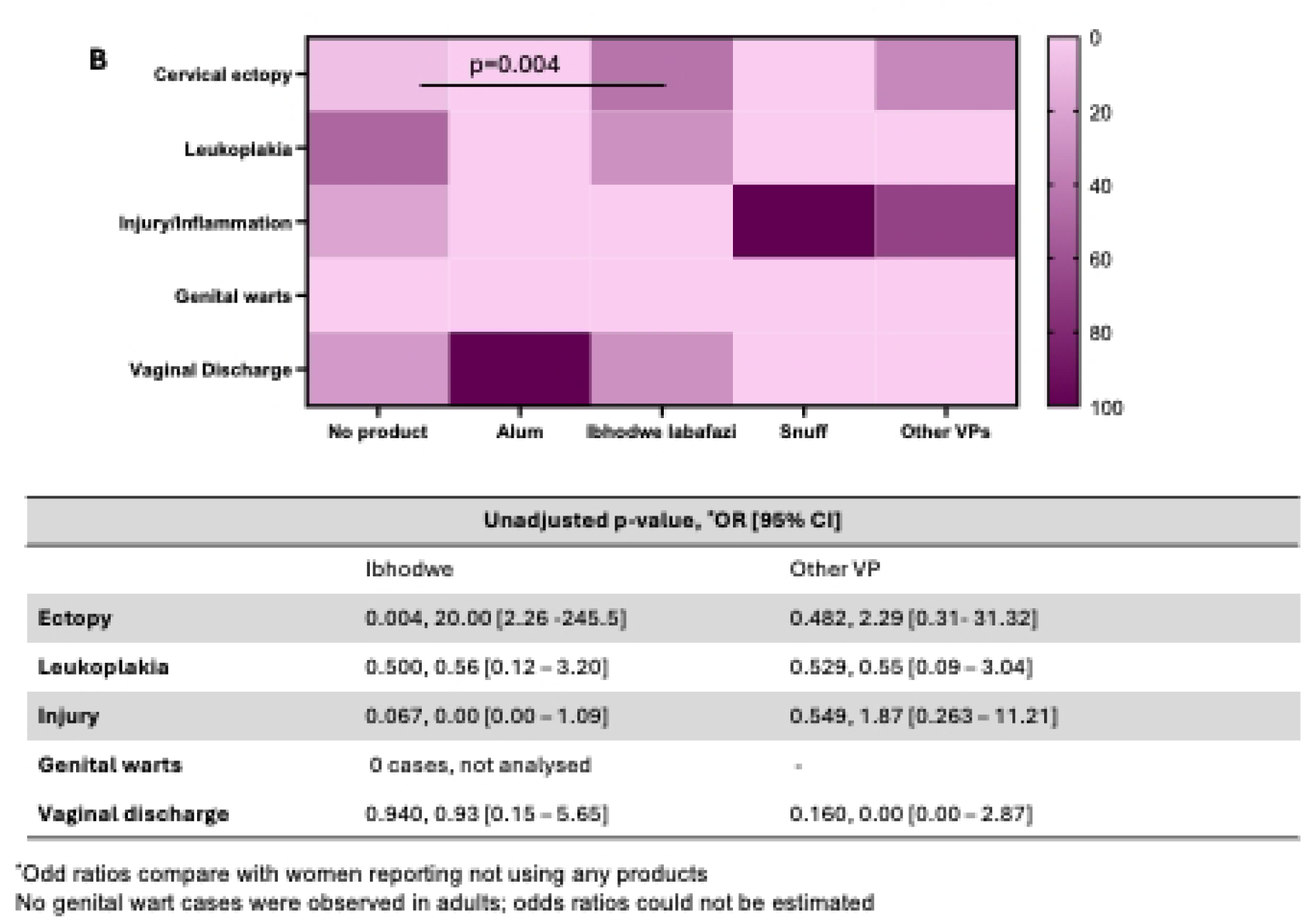
Cervicovaginal colposcopic findings among adolescents and adult women who use vaginal products compared to nonusers. **A.** Adolescents: Heatmap (top) shows frequencies of colposcopic findings by product use group—nonusers (n=63), Alum (n=27), Ibhodwe Labafazi (n=9), Snuff (n=6), and other vaginal products (Other VP, n=36). Table (bottom) shows unadjusted p-values (Chi-squared), odds ratios, and 95% confidence intervals comparing users vs. nonusers. **B.** Adults: Heatmap (top) for nonusers (n=19), Ibhodwe Labafazi users (n=9), and Other VP users (n=7). Table (bottom) shows corresponding unadjusted comparisons. Colour scale: blue indicates higher frequencies; pink indicates lower. p-values <0.05 considered significant. Heatmaps generated in GraphPad Prism v8.4.3.

Among adults, unadjusted analyses showed an association between ibhodwe labafazi use and cervical ectopy (OR 20.0; 95% CI 2.26–245.5; p = 0.004). This association did not persist after adjustment for potential confounders (aOR 0.94; 95% CI 0.26–3.89; p = 0.92; Figure 3B). In adjusted models, both intravaginal (aOR 3.7; 95% CI 1.2–11.8; p = 0.023) and ingested (aOR 2.9; 95% CI 1.1–8.5; p = 0.032) VSP use remained statistically associated with cervical ectopy. No specific products were associated with leukoplakia, inflammation, or abnormal discharge after adjustment.

### Prevalence of sexually transmitted infections and other infections amongst product users

Among adolescents, *Trichomonas vaginalis* infection was more prevalent among VSP users than non-users (p = 0.021; Fig 4A). No differences were observed for *Chlamydia trachomatis* or *Neisseria gonorrhoeae*. Among adults, overall STI prevalence was observed to be higher among users of ingested products (p = 0.001; Fig 4B); however, no statistically significant differences were observed when infections were examined individually. Product-specific infection analyses were limited by sample size.

**Fig 4.**
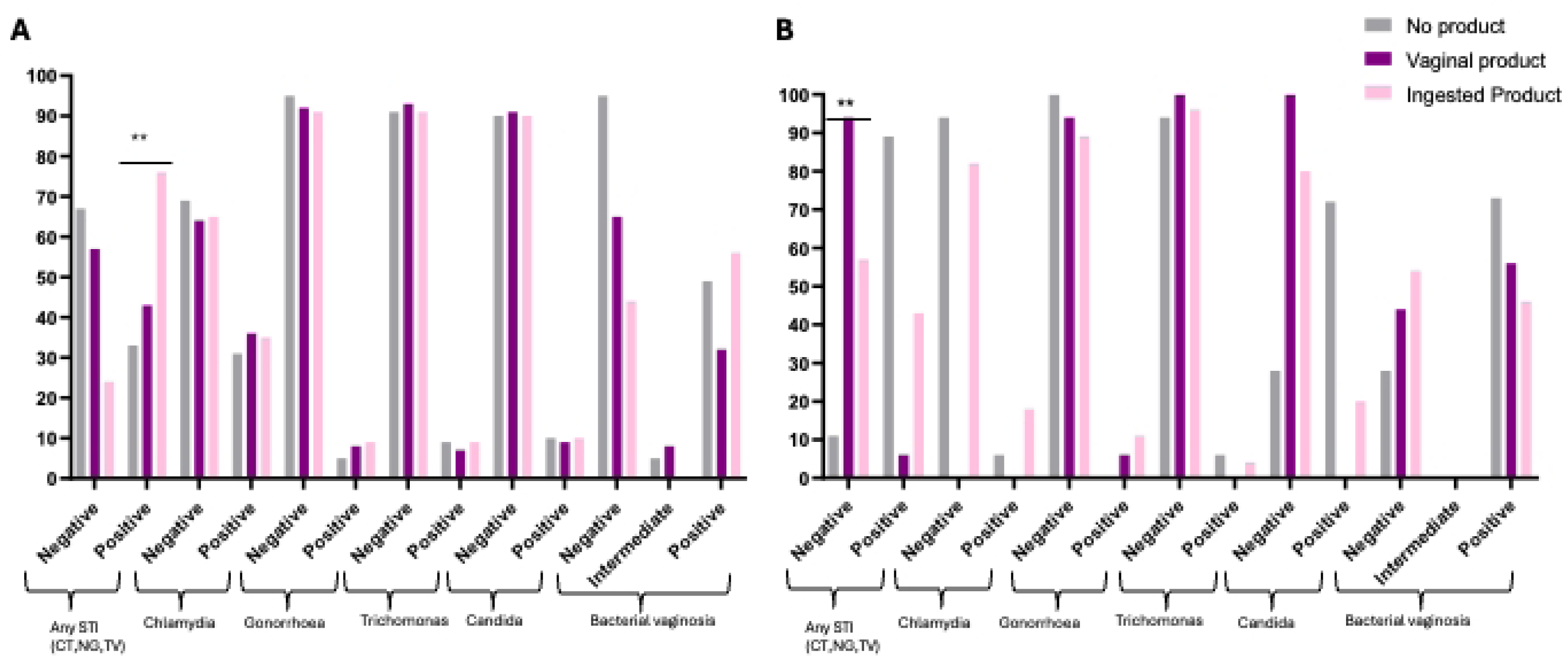
Bar plot of the prevalence of sexually transmitted infections and genital infections amongst product users. The prevalence of STIs (CT, TV, NG), BV, and Candida was measures amongst A) Adolescents. B) Adults. using vaginal/oral product. Fisher’s exact test was used to compare the prevalence of infection among product users (vaginally inserted (purple panel) / oral (light pink panel) in comparison to participants reporting to not use any on the products (no product: light grey) in the cohort and a p ≤0.05 was indicated by * and p≤0.001 was indicated by *** and were considered statistically significant.

### HPV prevalence (HR-HPV and LR-HPV) amongst product users by age

HPV genotype distribution varied by age group and product-use patterns (Fig 5; S3 Fig).

**Fig 5.**
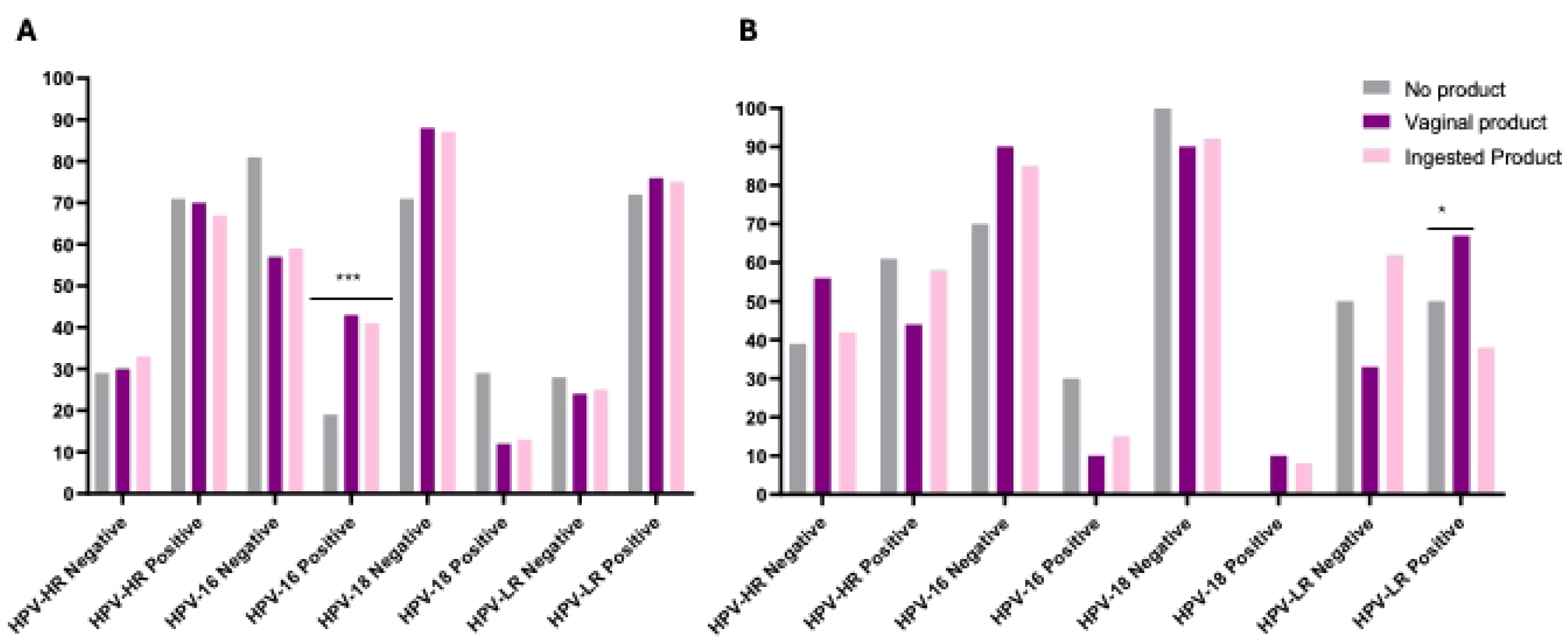
Prevalence of HPV infection by product-use group among adolescents and adults. The figure shows the prevalence of HPV infection among non-users (grey), vaginal product users (purple), and ingested product users (pink). Panel **A** presents data for adolescents, and Panel **B** presents data for adults. Within each panel, HPV infection prevalence is displayed for three categories: LR (low-risk HPV), HR (high-risk HPV), and HPV-16.

Among adolescents, high-risk HPV types 16 and 18 were most prevalent, with higher overall HR-HPV prevalence observed among both intravaginal and ingested VSP users, particularly HPV-16 (p = 0.007). Low-risk (LR) HPV types 62/81 and 6 were common but did not differ by product use. Among adults, LR-HPV types were more prevalent among vaginal product users (p = 0.005), while HPV-35 predominated among ingested product users (p = 0.001). Vaginal product users also had higher prevalence of HPV-62/81 (p = 0.005).

### Model performance and sensitivity analyses

Multivariable logistic regression models adjusted for age, hormonal contraceptive use, condom use, number of lifetime sexual partners, and vaginal pH showed no significant associations between general VSP use and cervicovaginal abnormalities among adolescents (adjusted ORs 0.6–1.2; all p > 0.05). These models were intended to estimate associations rather than predictive performance (S Index).

## Discussion

This study integrates behavioural assessment, colposcopic examination, and molecular diagnostics to characterise VSP use among AGYW in a high HIV/STI burden setting. By situating sociocultural practices within a biological framework, these findings contribute to emerging evidence that informal vaginal product use may intersect with mucosal health and infection vulnerability. Given ongoing global efforts toward cervical cancer elimination and STI reduction, understanding modifiable behavioural and structural exposures remains a public health priority. VSP use was highly prevalent, with substantial variability in product types. Alum use was predominantly reported by adolescents, whereas ibhodwe labafazi was more commonly reported by adults. These patterns are consistent with qualitative reports describing sociocultural norms related to vaginal cleanliness, tightness, and partner retention (12,13,23).

The observed association between VSP use and cervical ectopy among adults is noteworthy. Cervical ectopy is typically considered a physiological feature more common during adolescence and early reproductive maturation (24–26). The higher prevalence of ectopy among adult VSP users raises questions about potential relationships between product use and epithelial characteristics; however, temporality cannot be established and reverse causation remains possible. Similar epithelial alterations have been described in studies of some intravaginal cleansing agents (18,27), but direct mechanistic evidence specific to the products examined here remains limited.

The finding that alum use was statistically associated with lower odds of visible cervicovaginal injury among adolescents should be interpreted cautiously. Intravaginal practices are heterogeneous, and the astringent properties of alum may transiently alter epithelial appearance during colposcopic examination without necessarily reflecting improved mucosal integrity. Previous literature has also highlighted variability in biological effects across intravaginal products, with some appearing neutral and others associated with epithelial disruption (28). These observations underscore the importance of evaluating products individually rather than generalising across practices.

Exploratory infection analyses suggested a higher prevalence of *Trichomonas vaginalis* infection and HPV-16 among adolescent VSP users. These findings do not establish causality but are consistent with hypotheses proposing that intravaginal practices may influence susceptibility to certain infections through alterations in epithelial integrity or local mucosal immunity. Prior experimental work has demonstrated epithelial disruption and inflammatory cytokine responses following exposure to some intravaginal agents (18), although further longitudinal studies are required to clarify temporal relationships.

The differential distribution of HPV genotypes by age and product use also suggests potentially complex interactions between behavioural exposures, mucosal environment, and cumulative infection risk. However, given the modest sample size and cross-sectional design, these patterns should be considered hypothesis-generating rather than definitive.

### Study strengths and weaknesses

This study is among the few to combine behavioural assessment of VSP use with colposcopic examination, molecular HPV genotyping, and STI testing within a well-characterised cohort in a high HIV/STI burden setting. Age-stratified analyses allowed evaluation of associations within developmentally distinct cervical environments. Several limitations warrant emphasis. The cross-sectional baseline design precludes causal inference and does not establish temporality between VSP exposure and cervicovaginal outcomes. Self-reported product use may be subject to recall or social desirability bias, and the chemical composition of many products was unknown. Sample size, particularly among adults, limited statistical power for product-specific and genotype-specific analyses and contributed to wide confidence intervals. Multiple comparisons were conducted without formal correction, increasing the possibility of type I error. Additionally, the abstinence requirement prior to enrolment may have attenuated detection of acute mucosal changes. Longitudinal analyses of this cohort will be important to clarify temporal relationships between VSP exposure and cervicovaginal outcomes.

## Conclusion

VSP use was highly prevalent and demonstrated distinct age-specific patterns of product type and cervicovaginal findings. In this baseline cross-sectional analysis, VSP use was statistically associated with cervical ectopy among adults, while exploratory analyses suggested associations with certain STI and HPV patterns among adolescents. Although causality cannot be established, these findings highlight the importance of longitudinal investigation into the biological and behavioural pathways linking intravaginal practices with cervical health and infection risk, and may inform the development of culturally responsive sexual-health interventions.

## Data Availability

The data underlying the results presented in this study cannot be publicly shared because they contain sensitive sexual and reproductive health information collected from minors. Data may be made available upon reasonable request and with approval from the University of KwaZulu-Natal Biomedical Research Ethics Committee and the CAPRISA data access committee. Requests may be directed to the corresponding author

## Declarations

### Ethics approval and consent to participate

Ethics approval for this ancillary study was granted by the Biomedical Research Ethics Committee (BREC) sub-committee at the University of KwaZulu-Natal (BREC/00003306/ 2021).

### Consent for publication

All authors consent to the publication of this manuscript

## Authors’ contributions

**Conceptualization:** Passmore Jo-Ann, Jaspan Heather, Masson Lindi

**Data curation:** Radebe Phumla, Mkhize Pamela, Sibeko Singeziwe

**Formal analysis:** Radebe Phumla, Mkhize Pamela, Sibeko Singeziwe

**Investigation:** Radebe Phumla, Mntambo Ntombenhle, Ndlela Nonsikelelo, Liebenberg Lenine, Sibeko Singeziwe, Maphumulo Nokuthula, Ngcapu Sinaye, Samsunder Natasha, Humphries Hilton

**Methodology:** Passmore Jo-Ann, Jaspan Heather, Masson Lindi, Mkhize Pamela

**Project administration:** Mkhize Pamela

**Supervision:** Pamela Mkhize, Passmore Jo-Ann

**Visualization:** Radebe Phumla, Mkhize Pamela

**Writing – original draft:** Radebe Phumla, Mkhize Pamela, Sibeko Singeziwe

**Writing – review & editing:** All authors reviewed and approved the final manuscript

## Acknowledgements

We acknowledge the study participants and the clinical staff at CAPRISA Vulindlela Research Site for their contributions to this research. We also thank the University of KwaZulu-Natal and University of Cape Town for institutional support.

---

**S1 Fig.**
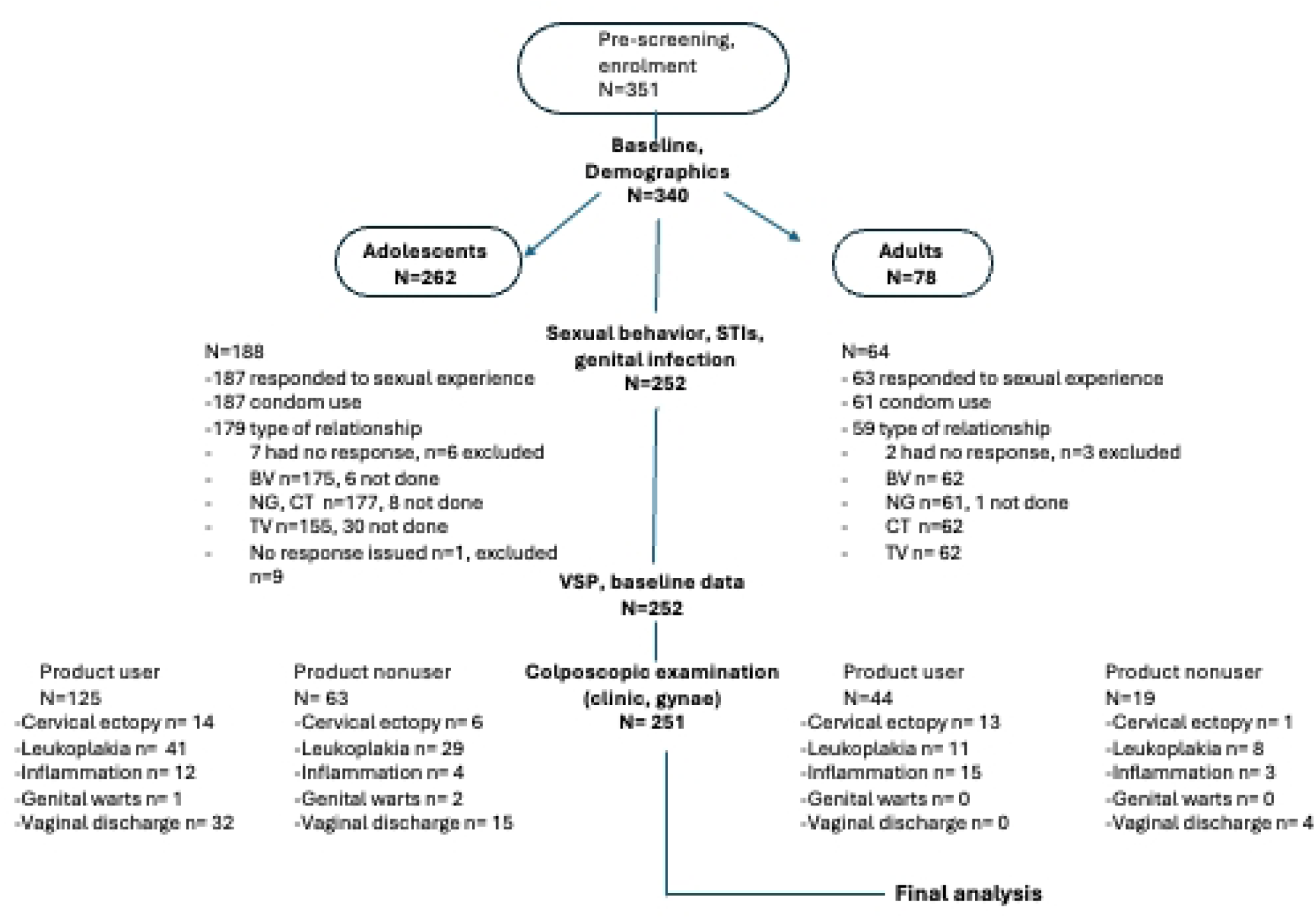
Study participant flow diagram. Flow of participants through the study from enrolment to final analysis. A total of 252 HIV-uninfected adolescent girls (14–19 years) and adult women (25–35 years) in KwaZulu-Natal, South Africa, were included in the baseline analysis. Participants were stratified by age group and vaginal-stimulating product (VSP) use status. Colposcopic assessment was performed for 238 participants. All 252 participants were included in the follow-up and final analysis.

**S2 Fig.**
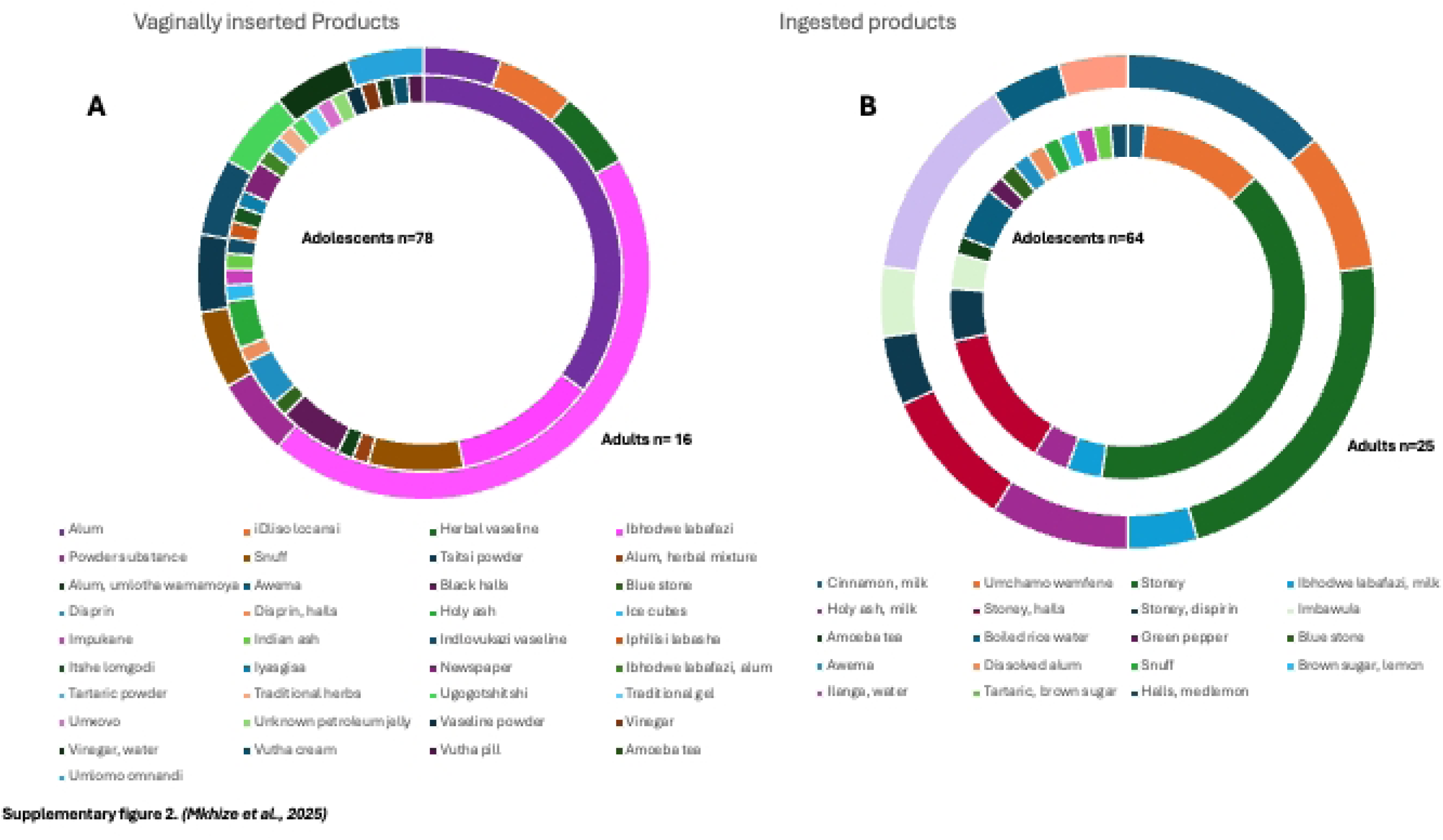
Distribution of product use in the cohort. Vaginal (left) and ingested (right) product use among adolescents (inner ring) and adults (outer ring) in KwaZulu-Natal, South Africa. Colours indicate specific products; some participants reported multiple types.

**S3 Fig.**
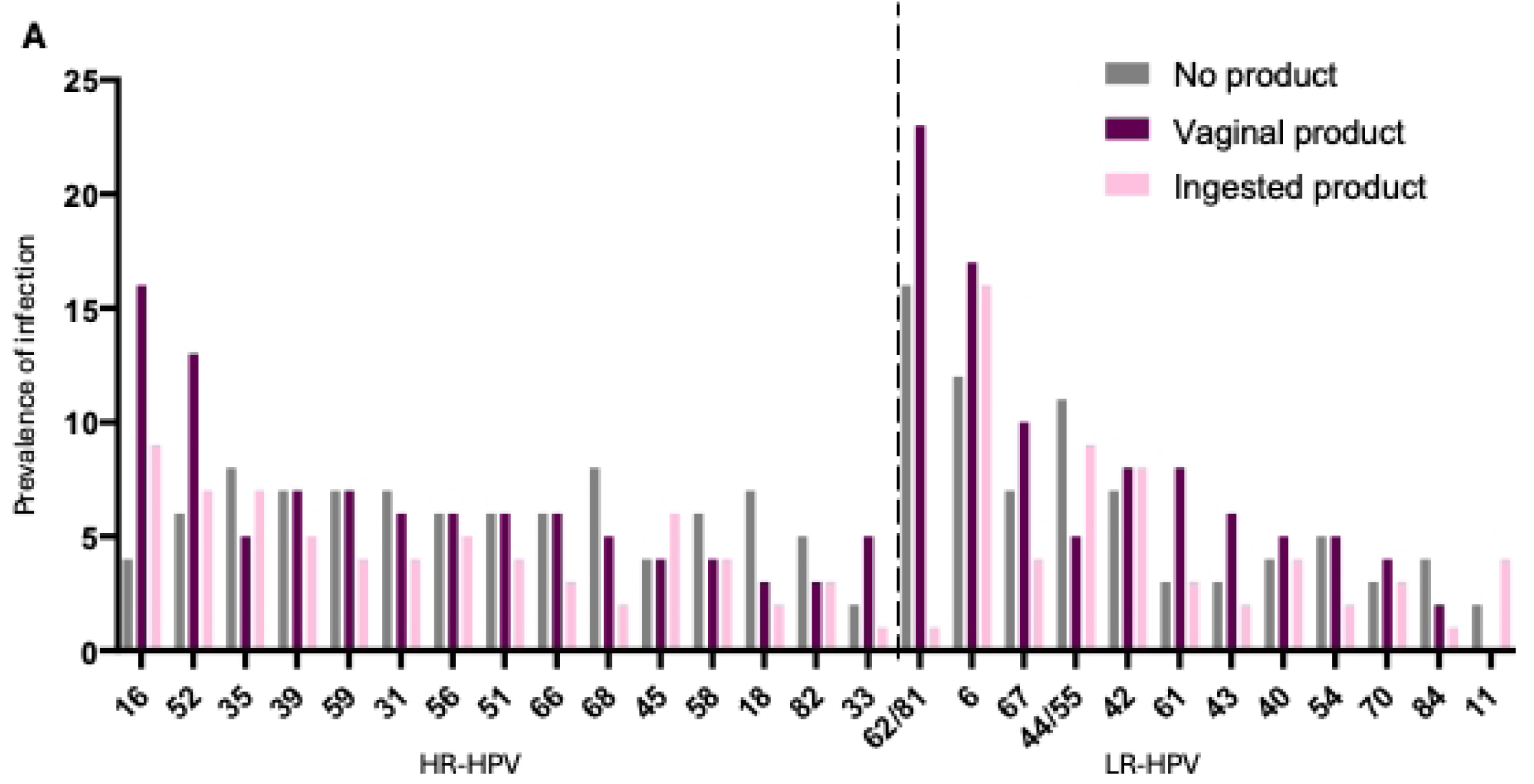
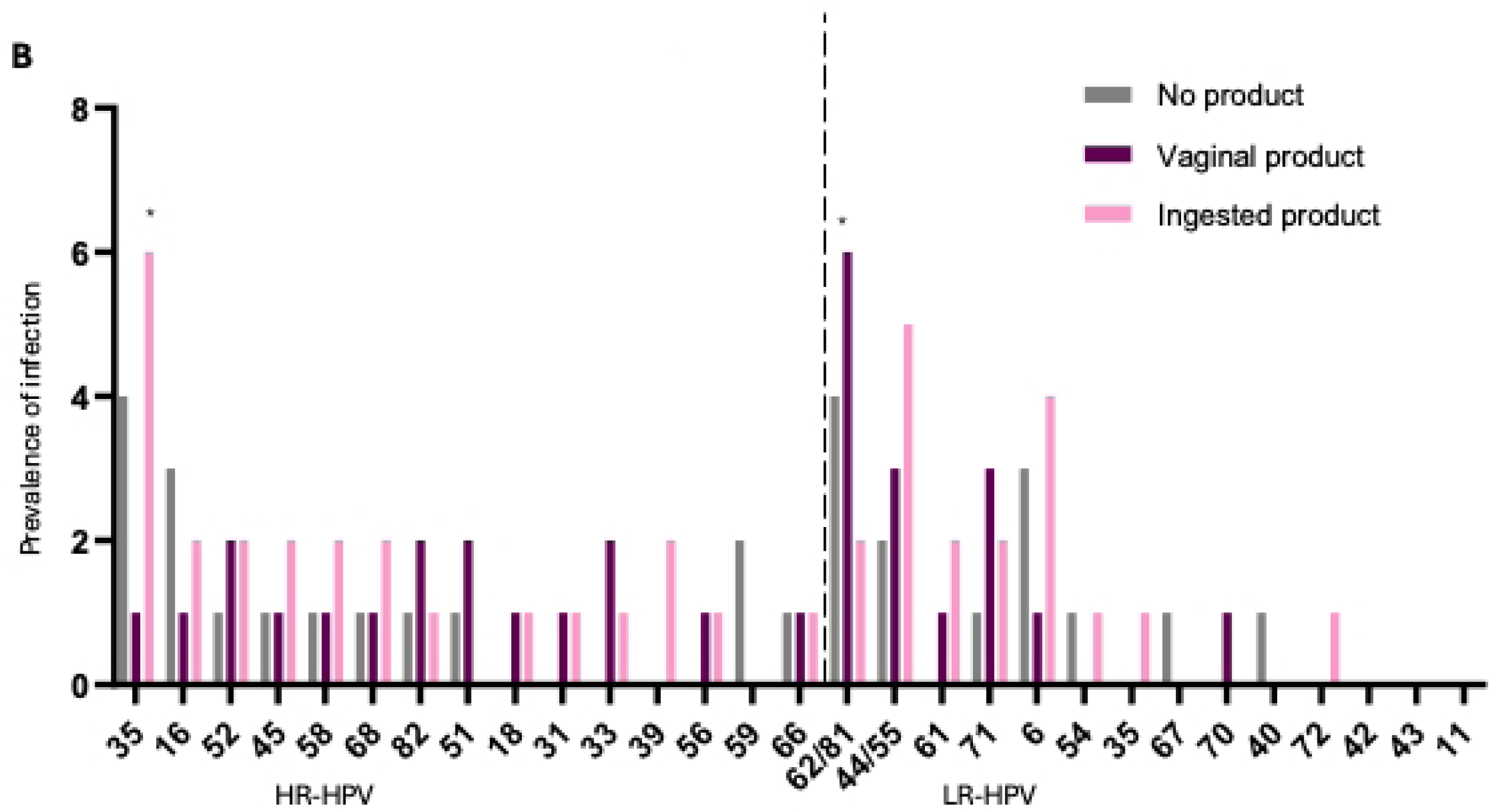
Clustered bar plot showing prevalence of high-risk (HR) and low-risk (LR) HPV subtypes by user groups. The clustered bar plot displays the distribution of HR-HPV and LR-HPV subtypes in adolescents (**A**, left panel) and adults (**B**, right panel), ranked by highest prevalence within each age group. User groups are shown in distinct colours, ingested user (pink), vaginal product user (purple) and nonuser (grey). A p-value <0.05 (*) was considered significant and were calculated using the Chi-squared test of Fishers pairwise test.

